# Etiological basis for chronic pain genetic variation in brain and dorsal root ganglia cell types

**DOI:** 10.1101/2025.07.03.25330832

**Authors:** Sylvanus Toikumo, Marc Parisien, Michael J. Leone, Chaitanya Srinivasan, Huasheng Yu, Asta Arendt-Tranholm, Úrzula Franco-Enzástiga, Christoph Hofstetter, Michele Curatolo, Wenqin Luo, Andreas R. Pfenning, Rebecca P. Seal, Rachel L. Kember, Theodore J. Price, Luda Diatchenko, Stephen G. Waxman, Henry R. Kranzler

## Abstract

Chronic pain is a complex clinical problem comprising multiple conditions that may share a common genetic profile. Genome-wide association studies (GWAS) have identified many risk loci whose cell-type context remains unclear. Here, we integrated GWAS data on chronic pain (*N* = 1,235,695) with single-cell RNA sequencing (scRNA-seq) data from human brain and dorsal root ganglia (hDRG), and single-cell chromatin accessibility data from human brain and mouse dorsal horn. Pain-associated variants were enriched in glutamatergic neurons; mainly in prefrontal cortex, hippocampal CA1-3, and amygdala. In hDRG, the hPEP.TRPV1/A1.2 neuronal subtype showed robust enrichment. Chromatin accessibility analyses revealed variant enrichment in excitatory and inhibitory neocortical neurons in brain and in midventral neurons and oligodendrocyte precursor cells in the mouse dorsal horn. Gene-level heritability in the brain highlighted roles for kinase activity, GABAergic synapses, axon guidance, and neuron projection development. In hDRG, implicated genes related to glutamatergic signaling and neuronal projection. In cervical DRG of patients with acute or chronic pain (*N* = 12), scRNA-seq data from neuronal or non-neuronal cells were enriched for chronic pain-associated genes (e.g., *EFNB2*, *GABBR1*, *NCAM1*, *SCN11A*). This cell-type-specific genetic architecture of chronic pain across central and peripheral nervous system circuits provides a foundation for targeted translational research.

## INTRODUCTION

Chronic pain is both a symptom and a primary disease. It affects about 20% of the global population and is associated with psychiatric and physical comorbidities^1^, contributing to high rates of morbidity and mortality^2^. Despite its substantial and growing impact on public health^3^, currently available treatments for chronic pain are nonspecific, of limited efficacy, and associated with substantial adverse effects, including addiction^4–6^. A deeper understanding of pain biology is crucial for the development of novel, targeted, safe, and effective treatments^7^.

Integration of genomic data into drug development pipelines can yield drug approval rates that are 2.6 times those that lack such information^8^ and could expedite the prioritization of new targets for chronic pain. Despite their diverse phenotypic manifestations, pain conditions exhibit high genetic overlap^9^, a common genetic risk profile^10,11^ and similar associated alterations of the central nervous system (CNS)^12,13^. Genome-wide association studies (GWAS) have revealed hundreds of genetic risk loci for chronic pain, which account for a substantial fraction of the trait’s heritability^14–16^. Previous GWAS of pain identified the brain as a tissue whose loci at expressed genes display enriched heritability^14,17,18^, and more so for chronic pain than acute pain^19^. However, despite the availability of transcriptomics data from various isolated brain regions, their cellular diversity remains unexplored.

Chronic pain can be caused and maintained by alterations in various components of the pain pathway, including dorsal root ganglia (DRG), the spinal cord dorsal horn^20,21^ and supraspinal brain centers^12,13^, with neuronal architecture organized into complex, hierarchically structured clusters of distinct cell types^22,23^. We recently showed that genetic variation that influences pain intensity is enriched in GABAergic neurons in the mid-brain^16^ and identified a role for mouse dorsal horn neuronal-subtype-specific open chromatin in multi-site chronic pain^24^. While these findings relate to individual pain phenotypes with substantial genetic overlap^16^, the shared downstream biological effects of the implicated genes on the complex cellular structure of the brain and dorsal horn are unknown.

We recently performed a large GWAS meta-analysis of chronic pain phenotypes (*N* = 1,235,695), that implicated 343 genomic loci and revealed pleiotropic associations with psychiatric disorders, immune traits, and brain structures^25^. Although this study revealed key genetic risk factors for chronic pain, the specific cell types that define the genetic associations underlying the cellular and circuit-based mechanisms^26,27^—key to therapeutic targeting^28,29^—are yet to be determined.

Here, we integrate multiple layers of single-cell omics from the brain, dorsal horn, and DRG with our GWAS meta-analysis of chronic pain to identify cell types and pathways relevant to the trait (Figure 1). This effort, made possible with datasets from two recent studies, advances our understanding of the physiological relevance of genomic associations for chronic pain. The first of these studies analyzed a comprehensive human brain transcriptomic dataset, using single-nucleus RNA sequencing (snRNAseq) of 3.369 million nuclei from 106 anatomical dissections across 10 brain regions^30^. The second study was of the neural basis of human somatosensation that used single-soma RNAseq data from human DRG (hDRG) neurons^31^.

**Figure 1.**
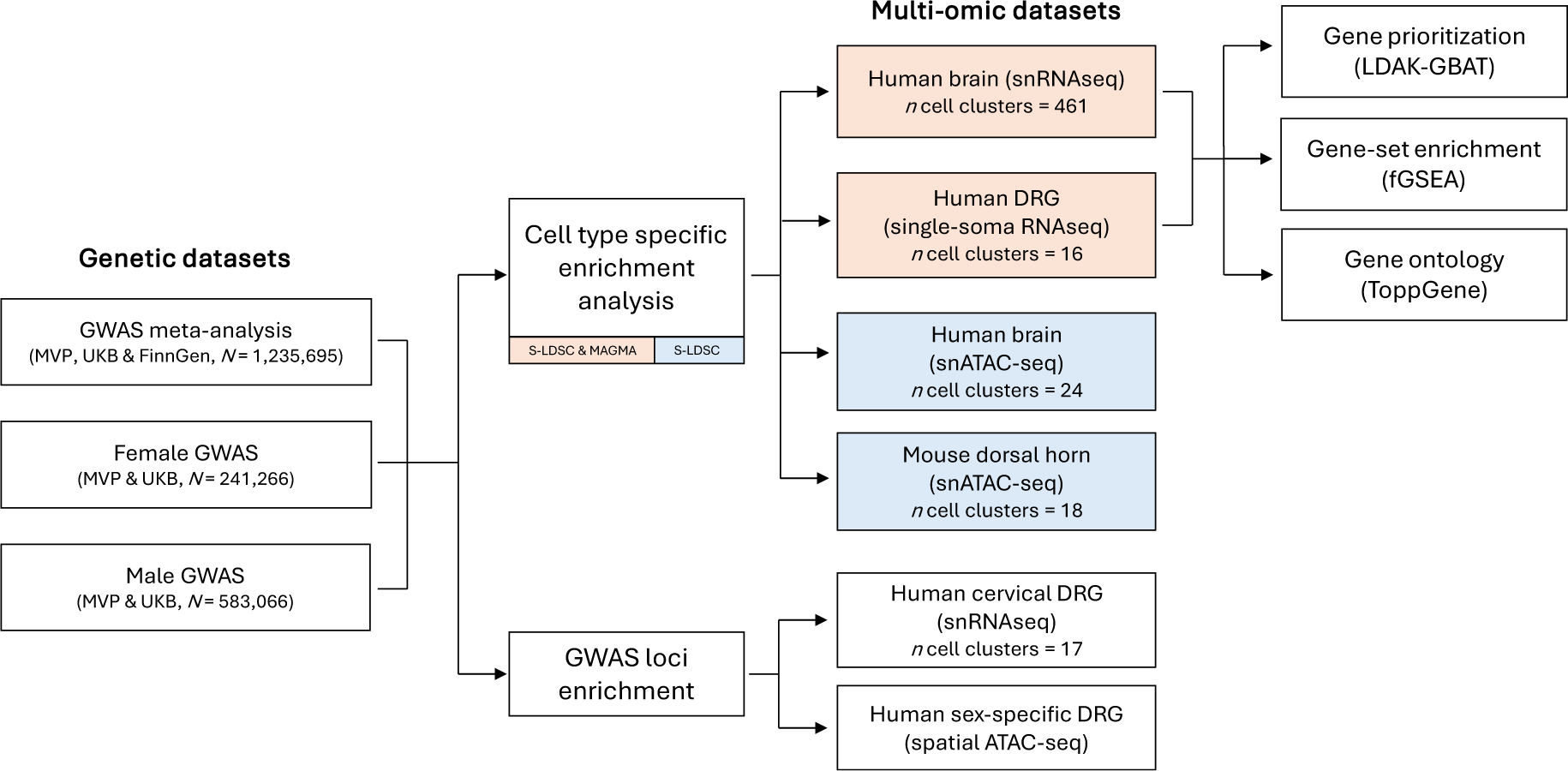
Study overview. Using S-LDSC, we first identified cell types enriched for chronic pain SNP-heritability across single-nucleus transcriptomic data from human brain and hDRG, as well as chromatin accessibility data from human brain and mouse dorsal horn. To test the robustness of cell type enrichment results in transcriptomic human brain and hDRG data, we used MAGMA, followed by gene prioritization and ontology analysis. Multi-omic datasets included in both S-LDSC and MAGMA analyses are shown in pink, while those included only in MAGMA are shown in blue. Finally, genetic loci from the pain meta-analysis were tested for enrichment in differentially expressed genes (snRNAseq) and sex-stratified GWAS for genes linked to differential chromatin accessibility (spatial ATAC-seq) from hDRG datasets.

## RESULTS

### Cell type identification using S-LDSC

The brain’s cellular diversity has a hierarchical structure, with a foundational classification into broad categories (excitatory neurons, inhibitory neurons, neuromodulatory neurons and non-neuronal cells) that extends to more intricate superclusters of cells subdivided into clusters and subclusters or cell types (Figure 2)^22,32^. Applying stratified linkage disequilibrium score regression (S-LDSC)^33^, we used this classification to assess chronic pain SNP heritability in 461 neuronal cell clusters generated from the extensive brain snRNAseq data^30^. The 461 cell clusters were further organized into the broad neuronal types of GABAergic (inhibitory, *n* = 131) and glutamatergic (excitatory, *n* = 209) subtypes, of which 15 (∼11.5%) and 76 (∼36.4%), respectively, were significantly associated with chronic pain at the false discovery rate (FDR) 5% level (Supplemental Table 1).

**Figure 2.**
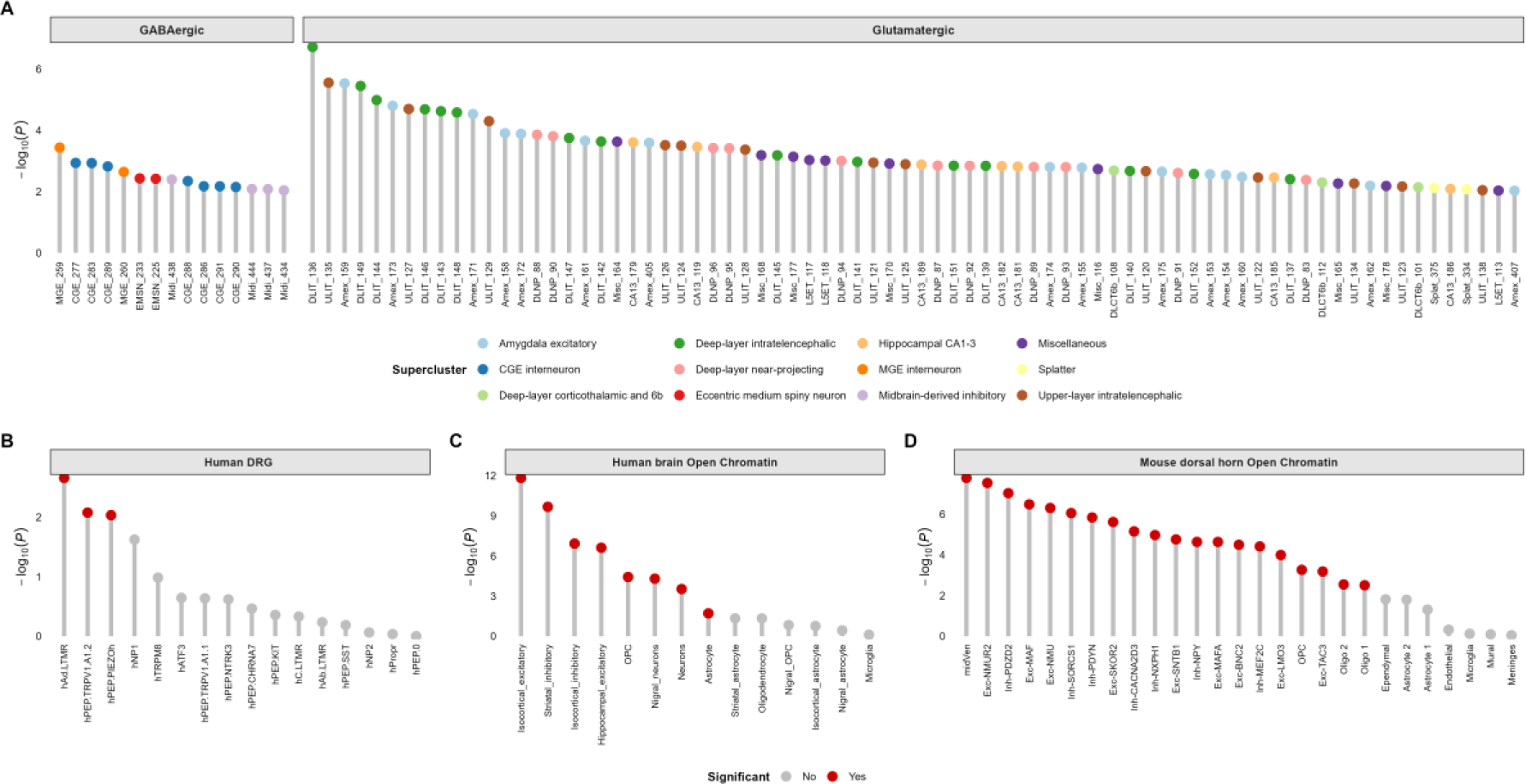
Cell type enrichments for chronic pain. Cell type numbers are denoted on the *x* axis, and statistical significance is on the *y* axis (−log_10_(*P*)). **(A)** The significantly enriched cell types for chronic pain (*n* = 91, FDR < 0.05) among 461 human brain cell clusters. Each cell type is colored based on its supercluster, and cell types are grouped into GABAergic (*n* = 15) or glutamatergic (*n* = 76) based on their neurotransmitter annotation. See Supplemental Table 1 for full results. **(B)** Human single-soma DRG cell type enrichment. **(C)** Human brain open chromatin cell type enrichment. **(D)** Mouse dorsal horn open chromatin enrichment. Cell type enrichments denoted by red in **B**-**D** are significantly enriched for chronic pain (FDR-corrected *p* < 0.05).

Hypergeometric tests of neurotransmitter annotations for the associated cell types revealed significant enrichment of glutamatergic—but not GABAergic—neurons in chronic pain heritability within the brain (glutamatergic *P* < 9 × 10^-18^; GABAergic *P* = 0.9). The enriched glutamatergic neurons include 25 intratelencephalic clusters and 11 deep layers from the prefrontal cortex; 11 amygdalar excitatory neuronal cell types; 7 hippocampal neuronal types annotated to subfield CA1-3; three cortex-wide annotations to deep layer 6b; and two “splatter” neuronal types, among others (Figure 2A, Supplemental Table 1). These enriched cell types were significantly over-represented in the prefrontal cortex (*P* < 4 × 10^-24^), amygdala (*P* < 2 × 10^-4^), and hippocampus (*P* < 0.01).

The entire body is innervated by DRG neurons, which serve as the primary origin of the pain pathway^34^. Thus, we explored peripheral neuronal cell types for chronic pain using single-soma hDRG data^31^. Of the 16 hDRG cell types analyzed, three (hAδ.LTMR, hPEP.TRPV1/A1.2, and hPEP.PIEZO^h^) show significant enrichment for chronic pain SNP heritability (FDR *p* < 0.05, Figure 2B, Supplemental Table 2).

Whereas studies support the enrichment of pain risk loci for mouse gene sets related to excitatory and inhibitory cellular pathways^35^, we examined whether the SNP heritability of human chronic pain shows similar cell-type enrichment. To do so, we used S-LDSC^33^ to partition the heritability of the chronic pain loci in putative cis-regulatory elements (CREs) from mouse dorsal horn neurons and glial cell types^24^ and human brain neurons and glial cell types^36^ (Methods). We found significant enrichment for pain among brain neurons aggregated across subtypes of excitatory (hippocampal and isocortical) and inhibitory (striatal and isocortical) neurons (Figure 2C, Supplemental Table 3). Of the brain glial cell types, the putative CREs of astrocytes that are found across the neocortex, hippocampus, striatum, and substantia nigra were significantly enriched in chronic pain-associated genetic variation (Figure 2C). Consistent with prior evidence^24^, we observed a broad enrichment of chronic pain variants across open chromatin of mouse spinal cord midventral neurons, inhibitory and excitatory neurons, and oligodendrocyte precursor cells (OPCs) (Figure 2D, Supplemental Table 4).

### Cell type identification using MAGMA

We used Multi-marker Analysis of GenoMic Annotation (MAGMA)^37^ to determine cell-type-specific enrichment of chronic pain gene-level associations. As expected, chronic pain variants mapped to the nearest gene were enriched largely in the brain (Figure 3A) and in neural subregions that process pain information^38,39^, such as the parietal, posterior insular, entorhinal, and medial frontal cortices, and the temporal gyrus (Figure 3B-C, Supplemental Table 5). We also observed robust enrichment in excitatory and inhibitory neurons, astrocytes, OPCs, and microglia (Figure 3D, Supplemental Table 6). Although immune cell types, including plasmacytoid dendritic cells, naïve B cells, and immune progenitor cells, were nominally enriched for chronic pain (*P* < 0.05), none survived multiple test correction (Supplemental Table 6).

**Figure 3.**
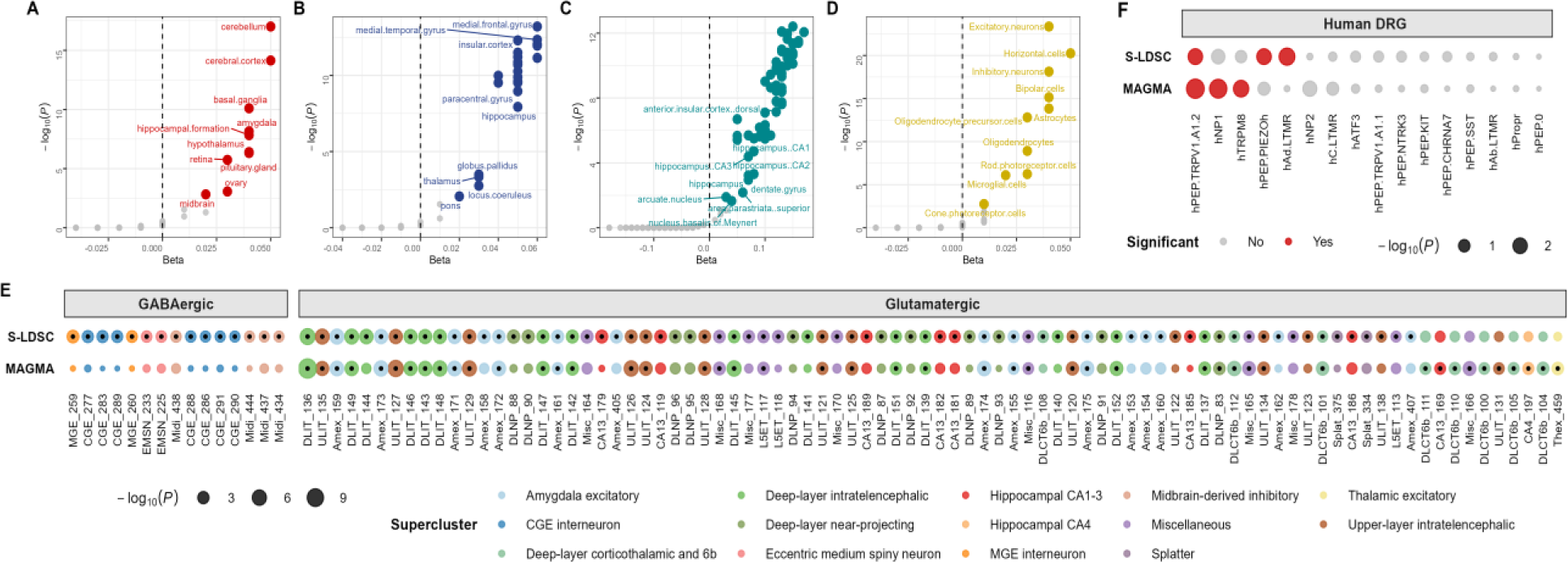
Comparison of MAGMA and S-LDSC cell type enrichment analysis. (**A–D**) MAGMA-based gene enrichment results. Significant heritability enrichments (FDR *p* < 0.05) are uniquely colored for each tissue- and cell-level analysis. Tissue-based enrichment analysis used data from **(A)** CONSENSUS, **(B)** FANTOM, and **(C)** Human Protein Atlas. **(D)** Cell-level enrichment analysis using RNAseq data from the Human Protein Atlas. **(E)** Combined MAGMA and S-LDSC results of significantly enriched cell types for chronic pain (101 unique cell types: 91 in S-LDSC vs. 47 in MAGMA, FDR *p* < 0.05) among 461 brain cell clusters. Each cell type is colored based on its supercluster, and cell types are grouped into GABAergic (*n* = 15) and glutamatergic (*n* = 86) based on their neurotransmitter annotation. Cell types that are significantly associated are marked by a black dot centered within each colored circle. In total, 37 cell types overlap across both methods. See Supplemental Tables 1 and 7 for full results. **(F)** Human single-soma DRG cell type enrichment. Cell type enrichments denoted by red in **F** are significantly enriched for chronic pain (FDR *p* < 0.05). In total, one cell type overlaps across both methods. See Supplemental Tables 2 and 8 for full results.

Consistent with the S-LDSC analysis, we also investigated the enrichment for chronic pain in 461 cell-type clusters from the brain snRNA-seq data^30^ and 16 cell types from single-soma RNAseq hDRG data^31^. Among the 461 brain cell-type clusters, only glutamatergic cell types (47 out of 109; 22.5%) were significantly associated with chronic pain (FDR *p* < 0.05), all of which showed strong enrichment (hypergeometric *P* < 9 × 10^-18^) (Figure 3E, Supplemental Table 7). The cell types with the most enrichments (*n* = 29) were predominantly intratelencephalic neurons from the prefrontal cortex (*P* < 3.8 *×* 10^-2^). Other significant cell types were excitatory, including seven cortex-wide annotations to deep layer 6b (*P* < 3.5 *×* 10^-2^); eight amygdalar neuronal cell types (*P* < 3.3 *×* 10^-2^); two hippocampal neuronal cell types (annotated to subfields CA1-3, *P* = 3.2 *×* 10^-2^ and CA4, *P* = 2.2 *×* 10^-2^); and one thalamic excitatory neuronal cell type (*P* = 9.5 *×* 10^-3^) (Figure 3E, Supplemental Table 7). Of the 16 hDRG cell types analyzed, we observed enrichment for hPEP.TRPV1/A1.2, hNP1, and hTRPM8 (Figure 3F, Supplemental Table 8).

Following prior studies^40^, we required that both our primary analytic method (S-LDSC) and the MAGMA gene property analysis provide strong evidence linking chronic pain GWAS associations to specific cell types in the brain and hDRG. These methods rely on different assumptions and modeling approaches: S-LDSC is SNP-based and evaluates GWAS heritability enrichment in the most cell-type-specific genes^33^, while MAGMA uses proximity-based SNP-to-gene mapping and tests whether gene-level associations increase linearly with cell type-specific expression^41^. To reduce false positives, we required both methods—each accounting differently for confounders such as gene size and linkage disequilibrium—to yield consistent results after multiple testing correction.

Using this approach, we found that glutamatergic neurons—across 37 cell-type clusters—are enriched in chronic pain, primarily in cortical interneurons, the hippocampus and the amygdala (Figure 3E). In the hDRG, both methods identified enrichment of the C-fiber thermoreceptor and nociceptor subtype hPEP.TRPV1/A1.2^31^ (Figure 3F).

### Contrasting cell-type enrichment for pain-related conditions

To test for the presence of cellular heterogeneity across pain conditions, we used S-LDSC to link brain and hDRG data with GWAS of other pain-related phenotypes, including knee pain, neck/shoulder pain, joint pain, low back pain, and migraine (Figure 4).

**Figure 4.**
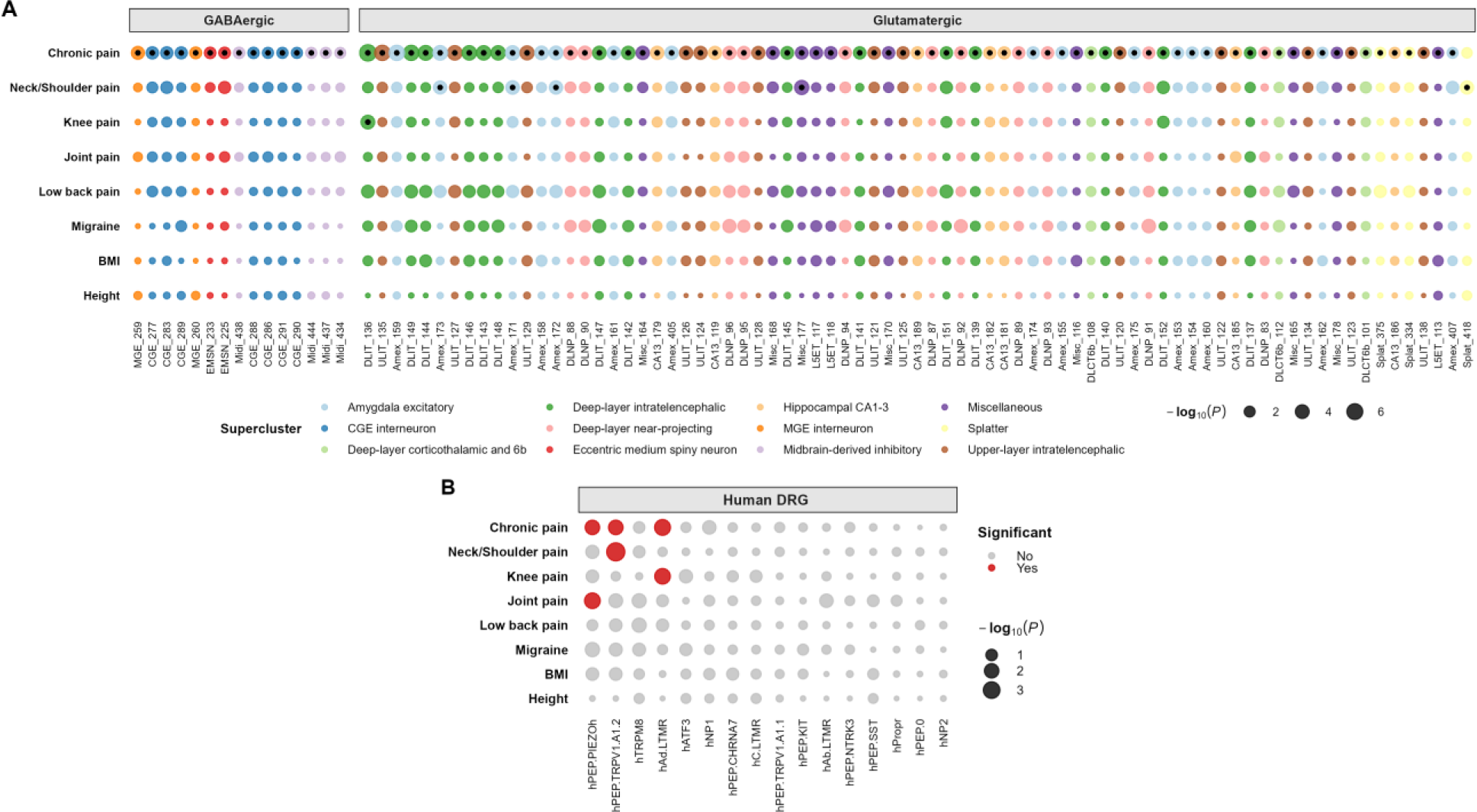
Cross-trait S-LDSC cell type enrichment analysis. **(A)** The significantly enriched cell types across traits (chronic pain, *n* = 91; Neck/shoulder pain, *n* = 5; Knee pain, *n* = 1; FDR *p* < 0.05) among 461 brain cell clusters. Each cell type is colored based on its supercluster, and cell types are grouped into GABAergic (*n* = 15) and glutamatergic (*n* = 77) based on their neurotransmitter annotation. Cell types that are significantly associated are marked by a black dot centered within each colored circle. In total, 4 significant brain cell clusters (DLIT_136, Amex_171, Amex_172, and Amex_172) overlap across traits. **(B)** Human single-soma DRG cell type enrichment. Cell type enrichments denoted by red in **B** are significantly enriched for chronic pain (FDR *p* < 0.05). All three significantly enriched hDRG cell types overlap in at least two traits. See Supplemental Tables 9 and 10 for full results.

The significantly enriched brain cell type for knee pain was an intratelencephalic glutamatergic neuron (DLIT_136, *P* = 4.3 *×* 10^-2^), consistent with previous findings of cortical glutamatergic neuronal activity in knee pain^42^. For neck/shoulder pain, five cell types showed significant enrichment, largely in amygdalar excitatory neuronal cell types (e.g., Amex_171, *P* = 3.1 *×* 10^-2^, Amex_172, *P* = 3.1 *×* 10^-2^, and Amex_173, *P* = 3.1 *×* 10^-2^). This finding is consistent with evidence that links amygdalar hyperactivity with neck/shoulder pain^28,43^. Most (5/6) cell types were also significantly enriched in the chronic pain meta-GWAS results, highlighting a potential cross-trait pleiotropic cellular etiology (Figure 4A, Supplemental Table 9). No cell types for joint pain, low back pain, or migraine survived multiple testing corrections (Supplemental Table 9).

In the hDRG, hPEP.TRPV1/A1.2 was significantly enriched for neck/shoulder pain (*P* = 2.5 *×* 10^-3^), consistent with the role of C-fiber thermoreceptors and nociceptors in pain sensation^31^. Joint pain was enriched for hPEP.PIEZO^h^ (*P* = 4.3 *×* 10^-2^), supporting a role for piezo mechanosensory channels^31,44^. The significantly enriched hDRG cell type for knee pain was hAδ.LTMR (*P* = 4.6 *×* 10^-2^), highlighting its potential role in tactile sensation and pain modulation in the context of knee pain^45^. Notably, all three cell types that showed significant enrichment across these traits were also enriched in S-LDSC results for the chronic pain meta-GWAS (Figure 4B). At FDR *p* < 0.05, no hDRG cell types were significantly enriched for low back pain or migraine (Figure 4B, Supplemental Table 10), which is expected for migraine since this is a disorder of the trigeminal ganglia (TG) system and there are important differences between the DRG and TG^46,47^.

### Sex-specific cell type enrichment for chronic pain

Sex differences could play a role in pain neurobiology^48,49^. There is emerging evidence of sex-specific enrichment of pain GWAS loci/genes in the human brain^18^ and DRG^49,50^, and mouse DRG^51^. However, the cell-type-specific context needed to clarify these enrichment patterns in the human brain and hDRG are unknown. We thus partitioned the heritability of sex-stratified pain GWAS loci^25^ in the brain and hDRG. In males, we identified significantly associated SNP heritability signals for chronic pain in 10 GABAergic and 65 glutamatergic (VGLUT1, VGLUT2, and VGLUT3) brain neuronal cell types. Looking at the broad neuronal types, glutamatergic neurons—but not GABAergic—were significantly enriched for chronic pain (hypergeometric *P* < 6 × 10^-17^). Among females, only the intratelencephalic glutamatergic cell cluster (DLIT_136, *P* = 1.1 *×* 10^-2^) was significantly associated with chronic pain SNP heritability (Supplemental Figure 1A, Supplemental Table 11).

In the hDRG, hPEP.PIEZO^h^, hNP1, and hTRPM8 were nominally significant (*P* < 0.05) cell types among females, and hPEP.TRPV1/A1.2, hAδ.LTMR, and hTRPM8 were nominally significant among males, though no cell type was significantly enriched after correction for multiple testing (Supplemental Figure 1B, Supplemental Table 12).

### Prioritizing heritable genes for chronic pain in the brain and DRG cell types

We extended our investigation using LDAK-GBAT^52^, fGSEA^53^, and ToppGene^54^ analyses to understand the biological mechanisms of chronic pain cell type enrichments. These analyses enabled us to estimate the chronic pain heritability attributable to individual genes (Supplemental Table 13), pinpoint heritable gene clusters in the brain and hDRG, and identify biological processes and pathways enriched for the gene clusters in each cell type.

Among the 37 glutamatergic neuronal types significantly enriched for chronic pain in the brain (Figure 3E), heritable gene clusters were enriched in 25 neuronal clusters (nominal *P* < 0.05), 10 of which were significant after multiple testing correction (FDR *p* < 0.05) (Figure 5A, Supplemental Table 14). Of the 94 heritable genes identified in the 10 cell types, 29 were shared among them (e.g., *ACTN1*, *BSN*, *DAG1*, *SLC45A4*, and *PTPRO*) (Figure 5B). In the hDRG, hPEP.PIEZO^h^, hNP1, and hTRPM8 were nominally significant (*P* < 0.05), though none were significant after correction for multiple testing (FDR *p* > 0.05) (Supplemental Table 15). Notably, 89/94 (∼94.7%) of the enriched genes in the brain cell types were also identified in hDRGs (Figure 5A and C).

**Figure 5.**
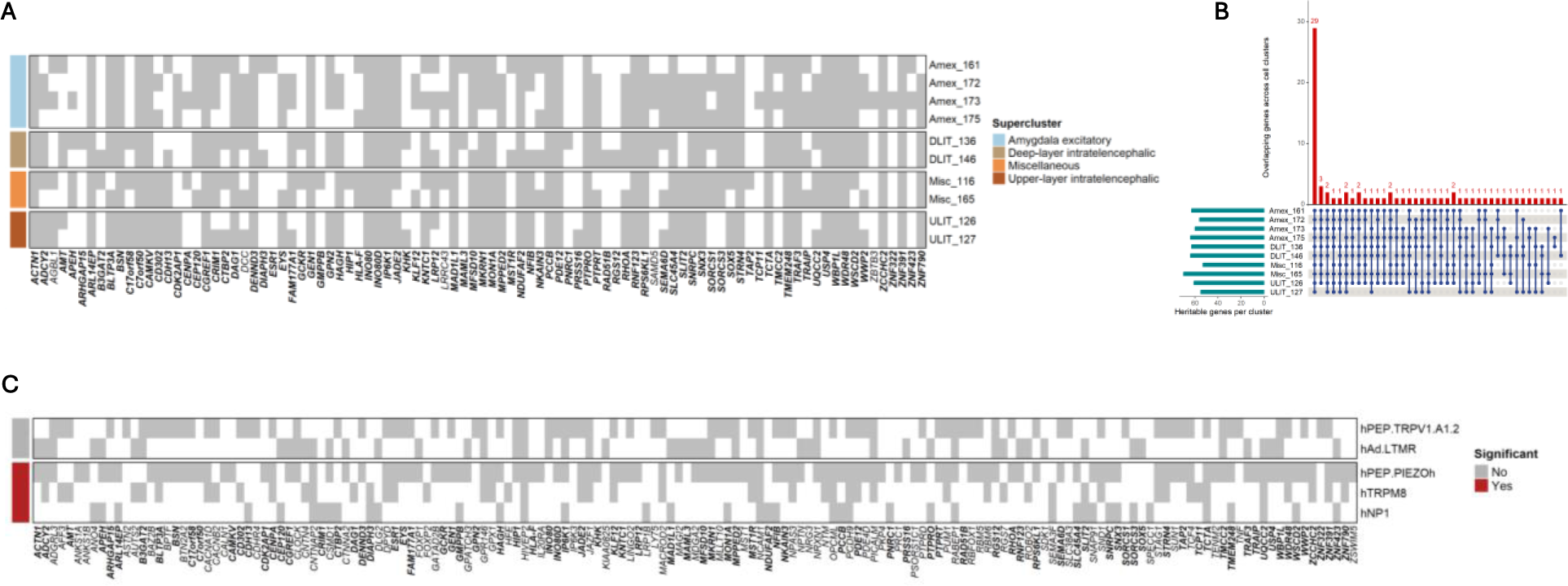
Heritable genes in brain and hDRG cell types. **(A)** The significantly heritable genes among 10 enriched brain cell clusters. Each cell type is colored based on its supercluster. **(B)** Gene overlap across brain cell types **(C)** The significantly heritable genes among 5 associated hDRG cell clusters. Overlapping genes in brain (A) and hDRG (C) are annotated in bold.

Gene over-representation analysis using heritable genes from the 10 significant brain cell types revealed enrichment for neuron projection, axon guidance, and synapse-related processes (Supplemental Figure 2, Supplemental Table 16). Particularly noteworthy is the intratelencephalic cell type ULIT_127, with heritable genes enriched for protein kinase activity (*PTPRO* and *PTPRT*), GABAergic synapse (*BSN*, *CDH13*, *CTBP2*, *DAG1*, and *PTPRO*), dorsal/ventral horn axon guidance (*DCC* and *SLIT2*), and neuron projection (*DCC*, *PTPRO*, *RHOA*, *SLIT2*, and *SEMA6D*) (Supplemental Table 16). In hDRG, prioritized heritable genes for hPEP.TRPV1/A1.2 were implicated in kinase activity (*IP6K3* and *IP6K1*), calcium channel activity (*CACNA1D* and *CACNB2*) glutamatergic synapse (e.g., *BSN*, *MDGA2*, *NPTN*, *NRXN1*, *PTPRD*), somatodendritic compartments (e.g., *ACTN1, BSN, CACNA1D, CNTNAP2, MAGI2, NCAM1*), and neuron projection (e.g., *AUTS2*, *CTNNA2*, *CNTNAP2*, *NCAM1*, *NRXN1*, *PTPRO*), among others (Supplemental Figure 2, Supplemental Table 17).

In summary, gene clusters with heritable links to chronic pain in the brain and hDRG cell types converge on biological processes related to synaptic function and neuron projection.

### Identification of differentially expressed or accessible genes linked to chronic pain risk

As in our current analyses, prior human pain GWAS studies that used S-LDSC and MAGMA analyzed transcriptomic data from healthy subjects or individuals of unknown disease status^14,15^. However, in rodents, hundreds of genes are differentially expressed in the DRG in response to painful stimulus^55^. At the same time, varying transcriptomic effects have been identified in the blood, spinal cord, and brain of humans experiencing pain^47,56^. Dysregulated genes are crucial for host immune defense and wound healing, as well as pain modulation, which enables the host to adopt survival behaviors^56,57^. We hypothesized that decoding the chronic pain GWAS at loci of pain-perturbed genes would explain a larger proportion of heritability than loci of homeostatic genes. To test the hypothesis, we selected differentially expressed genes (FDR *p* < 0.01) from a scRNA-seq dataset of cervical DRG samples from patients with acute or chronic pain^58^. The heritability enrichment at these genetic loci was assessed via QQ plots, where the gene-level MAGMA-derived chronic pain GWAS *p*-values were gauged against those expected by chance.

We found that differentially expressed genes in both neuronal and non-neuronal cell types exhibited steep slopes (λ) in these QQ plots (neuronal cells: λ=3.5 – 7.7; non-neuronal cells: λ=4.3 – 6.1), indicating greater enrichment of significant *p*-values for chronic pain than would be expected by chance (Figure 6A). Gene-set enrichment between the sets of genes differentially expressed in individuals with acute or chronic pain and the MAGMA pain-associated genes provide significant support for 526 genes, of which 88 were differentially expressed in at least two cell types (Supplemental Table 14). The top differentially expressed genes and those most strongly associated with findings from our chronic pain GWAS were *EFNB2* (*P*_MAGMA_=8.6 *×* 10^-7^), *GABBR1* (*P*_MAGMA_=6.2 *×* 10^-13^), *GRK4* (*P*_MAGMA_=6.2 *×* 10^-13^), *LSAMP* (*P*_MAGMA_=3.4 *×* 10^-34^), *NCAM1* (*P*_MAGMA_=2.5 *×* 10^-15^), *NRXN1* (*P*_MAGMA_=6.2 *×* 10^-13^), *PITPNM2* (*P*_MAGMA_=2.5 *×* 10^-15^), and *SEMA3F* (*P*_MAGMA_=2.5 *×* 10^-15^) (Figure 6B). We also found evidence of *SCN11A* loci enrichment in a neuronal cell cluster (*P*_MAGMA_=6.8 *×* 10^-4^, Supplemental Table 18). We observed an overlap between differentially expressed genes in cervical DRGs and heritable genes from the brain (27/95)^30^ and pain-free hDRGs (58/164)^31^ (Figure 6C).

**Figure 6.**
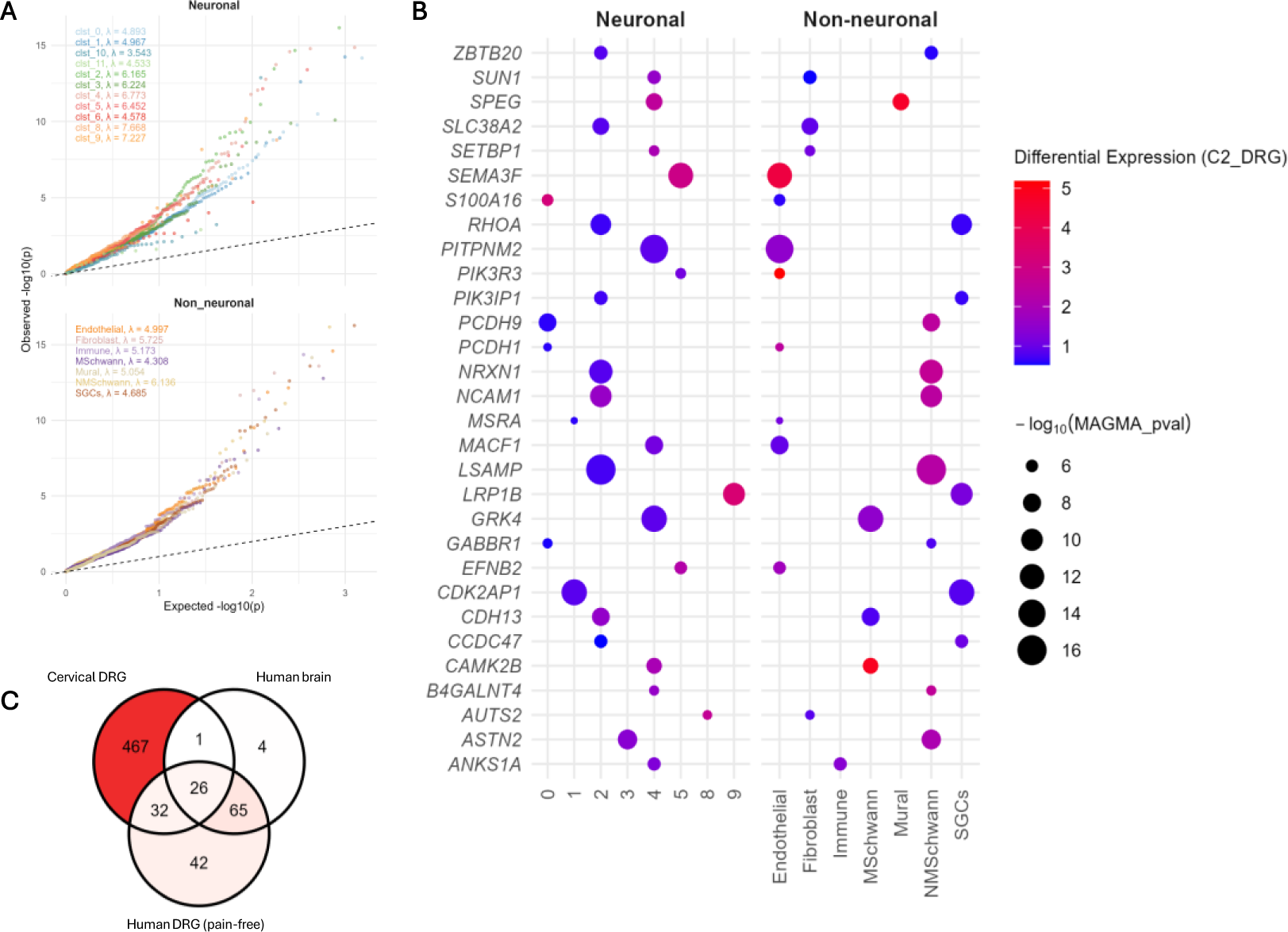
Chronic pain associated genes in cervical DRG of patients with acute or chronic pain. **(A)** QQ plots of pain-associated genes in neuronal and non-neuronal cervical DRGs. **(B)** The top 30 differentially expressed genes in neuronal and non-neuronal cervical DRG cell types that are associated with chronic pain risk. **(C)** Gene overlaps between cervical DRGs and gene-level results from brain and hDRG (pain-free) cell types.

Using the same approach as above, we also tested for sex-specific enrichment of GWAS loci in differentially accessible regions (DARs) from a spatial ATAC-seq hDRG dataset comparing neuronal chromatin accessibility between sexes^59^ (females: λ=2.5; males: λ=3.4; FDR *p* < 0.01; Supplemental Figure 3A). In females, hDRG DAR–gene associations included genes encoding the netrin-1 receptor *DCC* and sodium transporter *SLC4A10* (Supplemental Figure 3B, Supplemental Table 19). In males, the top hDRG DAR genes most strongly enriched for chronic pain genetic signals were involved in kinase activity (*CAMKV*, *DCAKD*), calcium signaling (*ERBB3*, *ITPR3*), and synaptic function (*BSN*) (Supplemental Figure 3B, Supplemental Table 19).

## DISCUSSION

In this study, we investigated the cell-type context of genetic associations for chronic pain in the central and peripheral nervous systems. By integrating GWAS and single-cell omics data, we identified enrichments of glutamatergic neurons in the brain, and peripheral enrichments for hDRGs in C-fiber neuronal subtype hPEP.TRPV1/A1.2. Analysis of chromatin accessibility revealed enrichment for pain in neocortical inhibitory and excitatory neurons in the human brain, and midventral neurons, inhibitory and excitatory neurons, and OPCs in the mouse dorsal horn.

Analysis of brain snRNAseq data using a broader chronic pain GWAS meta-analysis— which includes our earlier pain intensity GWAS—reveals predominant enrichment in glutamatergic neurons, augmenting prior findings of GABAergic enrichment^16^. Greater support for the role of glutamate here may reflect the greater resolution of our current dataset, which spans 461 cell types, substantially more than the 25 cell types previously assessed, potentially revealing new biological patterns. Dysfunction in the metabolism of GABA and glutamate, resulting in varying profiles of their metabolites in brain, has been observed across different pain conditions^60,61^. Studies in mice have also shown suppression of glutamatergic activity in the ventral tegmental area (VTA), with enhanced VTA glutamatergic inputs to the prelimbic cortex, which diminish pain-like behaviors^62^.

Lower medial prefrontal cortical glutamate levels have also been seen in people with chronic pain^63^. However, the genomic contribution of chronic pain variants to glutamatergic signaling in humans has been largely unsubstantiated. Thus, our finding that genes involved in chronic pain are enriched in intratelencephalic, hippocampal, and amygdalar excitatory neurons that express glutamatergic transporters (VGLUT1, VGLUT2, and VGLUT3) provides genomic insights into the molecular identity of key neuronal subtypes in the brain, consistent with aspects of the mesolimbic theory of pain^64^. These observations of gene enrichment across CNS regions—including the cortex, hippocampus, and amygdala—may have important implications for pain therapeutics, highlighting the need for analgesic drugs that cross the blood-brain barrier into the CNS, a consideration that may be overlooked in early-stage drug development^65^.

In the periphery, glutamatergic pain-signaling neurons detect noxious stimuli and transmit fast, sharp pain signals via Aδ-fibers and slow, dull pain via C-fibers^29^. These first-order pseudounipolar neurons, with cell bodies in the DRG, synapse via glutamate on second-order neurons in the spinal cord, before signals are relayed to third-order neurons in the brain regions that process pain^66^. Musculoskeletal pain heritability has been mapped to both C-fiber peptidergic (PEP1) and non-peptidergic (NP2) neuronal types in the macaque DRG^67^. Extending these findings to humans, we show that chronic pain genetic associations are enriched in the C-fiber peptidergic subtype hPEP.TRPV1/A1.2 in hDRGs. Different DRG neuronal types may be enriched for pain in different bodily sites^68,69^. In macaques, PEP1 neurons are enriched for facial, neck/shoulder, stomach and hip pain, and NP2 neurons for back and hip pain^67^. Here, we observed that genetic signals contributing to joint, knee and neck/shoulder pain are enriched in hPEP.PIEZO^h^, hAδ.LTMR, and hPEP.TRPV1/A1.2, respectively. Taken together, our results differentiate the DRG neurons that mediate genetic risk for pain in humans, implicating A-fiber mechanosensory channels, C-fiber thermo-nociceptors and tactile sensation neurons^31^.

Gene-level heritability and over-representation analysis in brain and hDRGs converge to show that neuronal development and differentiation, and regulation of synaptic organization are key mechanisms of chronic pain. In cervical DRGs from chronic pain patients^58^, several chronic pain–associated genes (e.g., *EFNB2*, *GABBR1*, *GRK4*, *NCAM1*, and *NRXN1*) were differentially expressed across both neuronal and non-neuronal cell types. In contrast, differential expression of *SCN11A* was specific to neurons. These findings highlight the key molecular features of human sensory neurons across the peripheral nervous system.

As gene expression profiles are largely determined by distinct epigenomic signatures at gene promoters and enhancers, we explored enrichment for pain using single-nuclei ATAC-seq (snATAC-seq) data from the human brain and mouse dorsal horn. We identified putative cis-regulatory elements in brain excitatory (hippocampal, isocortical) and inhibitory (striatal, isocortical) neurons, and astrocytes. In line with our recent work^24^, we also found that chromatin accessibility in the spinal cord, involving superficial dorsal and midventral neurons, oligodendrocytes and OPCs of the mouse dorsal horn was enriched for chronic pain. These findings shed light on the neural gene regulatory circuits and provide a framework for interpreting non-coding genomic variants linked to chronic pain.

Our findings build upon previous studies that reported sex-specific tissue enrichment of pain-associated signals in the human brain^18^ and DRG^49,50^, and mouse DRGs^51^. Specifically, we characterized sex-related cell-type enrichment patterns for chronic pain and showed that glutamatergic (VGLUT1, VGLUT2, and VGLUT3) neuronal cell types were enriched among males, while only an intratelencephalic glutamatergic neuron was enriched in females. Extending our analysis to sex-specific chromatin accessibility in hDRG neurons, we found that female DAR-gene associations involved *DCC* and *SLC4A10*, while male-enriched genes were linked to kinase activity (*CAMKV*, *DCAKD*), calcium signaling (*ERBB3*, *ITPR3*), and synaptic function (*BSN*).

Our findings would be enhanced by the availability of additional information at several levels. First, the brain snRNA-seq dataset^30^, which comprises 461 cell-type clusters from 10 brain regions, although the most comprehensive to date, is incomplete. Other brain regions involved in nociceptive signaling, such as the VTA^62^ and midbrain periaqueductal gray^70^, are not represented in the snRNAseq data. Second, snRNAseq and related omics datasets do not include individuals of non-European ancestry, limiting our ability to generalize the findings to a more diverse population. Furthermore, the greater brain cell-type enrichment for pain observed in males than females may be attributable to the predominance of males in the GWAS, primarily driven by the Million Veteran Program cohort. GWAS meta-analyses of chronic pain in large samples that are balanced on sex and ancestrally diverse are needed.

Gene-based analysis revealed differential expression of *SCN11A* in cervical DRG neurons, but not of *SCN9A* or *SCN10A*. Given the established genetic links between *SCN9A* and *SCN10A* and pain^71–73^, along with mechanistic evidence implicating Nav1.7-SCN9A, Nav1.8-SCN10A, Nav1.9-SCN11A as key mediators of DRG neuron-driven pain signaling^74^, the absence of association signals for *SCN9A* and *SCN10A* was unexpected. This discrepancy may reflect the strong contribution of Nav1.9-SCN11A to the control of excitability of both normal DRG neurons^75–77^ and in disease states caused by channel mutations^73^ or inflammation^78^. Alternatively, it may reflect phenotypic dilution in the GWAS meta-analysis, which included relatively few true cases of neuropathic pain. A more targeted GWAS using a physiologically relevant phenotype—such as painful peripheral neuropathy—may capture more fully the genetic contributions to neuropathic pain and hDRG function. Another critical advancement would be to conduct the same analyses using developmental datasets^79,80^ to gauge the contribution of chronic pain genetic variation in human brain and DRG development.

Transcriptomic data from disease-free individuals has been useful for identifying disease-relevant tissues and cell types^81^, but bulk tissue sequencing can miss signals specific to individual cell types. Single-cell sequencing overcomes this limitation, has been successfully applied to psychiatric disorders^82,83^, and was extensively used in our study. However, data from healthy individuals alone provide an incomplete view, as only comparisons between diseased and healthy states can reveal genes that are differentially regulated in pain-related pathways. The greater number of heritable genes identified in chronic versus acute pain in the cervical DRG snRNA-seq data supports this point (Figure 6C). The field is ripe for large-scale single-cell data collection across the pain matrix (nerves, DRG, spinal cord, brain) to identify pain-perturbed genes and enable high-resolution decoding of chronic pain GWAS findings.

In conclusion, our findings extend prior work by mapping genomic loci associated with chronic pain onto human brain and sensory neuronal types to identify its cellular origin. Genetic influences on pain identified in this study are predominantly enriched in glutamatergic neurons within the brain, and in C-fibers of the hDRG. Heritable genes for pain in both cell types converge on synaptic function and neuronal development. Combined with gene enrichment analysis in cervical DRGs that revealed enrichment of differentially expressed genes in neuronal and non-neuronal cell types, this study defines a broad set of central and peripheral neuronal features for pain genetic association, offering a basis for translational studies.

## METHODS

### GWAS data selection

We included summary statistics from the largest GWAS meta-analysis to date of chronic pain (*N* = 1,235,695 individuals of European-like ancestry (EUR))^25^. The meta-analysis combined GWASs from three biobanks. The first GWAS was performed by us in the Million Veteran Program (MVP; *n* = 436,683 EUR individuals)^16^. This study examined pain intensity (as a proxy for chronic pain), represented by the median of the annual median pain ratings measured across multiple years with an 11-point ordinal numeric rating scale, a consistent and valid measure of self-reported pain^84,85^. The second GWAS was conducted in the UK Biobank cohort (UKB, *n* = 387,649 EUR individuals)^14^. That study examined multisite chronic pain (MCP), defined as the count of chronic pain reported across 7 bodily sites (i.e., head, face, neck/shoulder, back, stomach/abdomen, hip, knee), which yields an 8-point ordinal score. The third GWAS was of “pain (limb, back, neck, head abdominally)” ascertained using ICD-9 and ICD-10 codes (Finnish version) from the FinnGen cohort (data freeze 10; *n* = 411,363 EUR individuals)^86^. The FinnGen study assessed overall chronic pain, with cases (*n* = 189,683) required to have one or more ICD-9 or −10 diagnostic codes for 16 disorders with a pain component (occurring in joints, limbs, neck, head, abdomen, and back) and none for controls (*n* = 221,680).

In addition to the chronic pain meta-GWAS, we selected seven comparison phenotypes that were both polygenic and had been studied in large GWASs. These phenotypes were selected to address concerns about specificity and phenotypic/genetic pleiotropy across pain phenotypes. We used published GWAS data for knee pain (UKB only, *n* = 254,380)^10^, neck/shoulder pain (UKB only, *n* = 237,825)^10^, joint pain (UKB and FinnGen, *n* = 722,279), low back pain (UKB and FinnGen, *n* = 693,056), and migraine (UKB and FinnGen, *n* = 802,589)^86^. We also included two phenotypes—height (*n* = 4,080,687)^87^ and body mass index (BMI) (*n* = 339,224)^88^—as negative controls, as their genetic basis differs substantially from pain-related disorders and is not rooted in the CNS.

For sex-stratified analyses, we included summary statistics from our GWAS meta-analysis of chronic pain among 583,066 males and 241,266 females of European ancestry^25^. The meta-analysis combined sex-stratified GWASs of MVP pain intensity (male; *n* = 404,510 and female; *n* = 32,173)^16^ and UKB MCP (male; *n* = 178,556 and female; *n* = 209,093)^51^.

### Multi-omic data selection

To establish a broad biological enrichment pattern of the genetic associations for chronic pain, we downloaded bulk and single-cell RNAseq data from six human tissues and cell types from the Human Protein Atlas (v.24)^89^. These datasets comprise 1) 50 human tissues from GTEx v.8^90^ and the Human Protein Atlas (v.24)^89^; 2) 46 tissues from the FANTOM consortium^91^; 3) 193 brain subregions from the Human Brain Tissue Bank of Semmelweis University^89,92^; 4) single-cell RNA-seq data from 81 cell types in the Human Protein Atlas (v.24)^89^; 5) 19 immune cell types from the Human Protein Atlas (v.24)^89^ and 6) 30 immune cell types from Monaco et al^93^. For data processing, gene expression levels measured in transcripts per Million (tpM) were log-transformed (as is typical for such data, we used log_2_(1+tpM)) to compress the scale and reduce outliers, with the mean-transformed expression for each gene in each cell type retained for further analysis.

To measure cell type enrichment of chronic pain in the brain and dorsal horn with high sensitivity, we obtained 1) snRNAseq data from a study of the transcriptomic diversity of cell types in approximately 100 locations across the human brain, which identified 461 cell-type clusters and 3313 subclusters^30^, and 2) single-soma RNA seq data from a study of human somatosensory physiology using hDRG, which identified 16 cell-type clusters^31^. Both datasets are the most comprehensive human brain and hDRG snRNAseq data available to date.

To examine chromatin accessibility enrichment patterns for chronic pain, we obtained snATAC-seq data from a study of the human brain: 10 samples spanning isocortex (*n* = 3), striatum (*n* = 3), hippocampus (*n* = 2) and substantia nigra (*n* = 2), and 70,631 total cells^36^. We also obtained snATAC-seq data from a study of the mouse dorsal horn, that harvested single nuclei from lumbar segments L3-L5 from 10 mice (5 female/5 male, 7-8 weeks old), pooled them and identified 74,437 glia and 19,073 neurons^24^. Neurons were grouped into 18 species conserved subtypes^24^.

### Calculation of cell-type expression specificity

We created a gene expression specificity matrix for each cell type by regressing expression levels of gene features with cell-type specificity using a binary measure (examining whether the cell type is present [yes=1] or absent [no=0]) among the defined cell clusters, adjusted for the age and sex of sample donors. For each cluster, genes were ordered by decreasing association test statistics (using gene expression as a marker of the cell type), with the top 1000 retained for S-LDSC analyses^33^.

### Heritability enrichment of chronic pain risk loci

Using S-LDSC^33^, we partitioned the SNP heritability for chronic pain and examined the enrichment of the partitioned heritability both in snRNAseq data from the adult human brain^30^ and single-soma RNAseq data from the hDRG^31^. Following a previously described LDSC pipeline^94^, we also estimated conditional heritability enrichment of chronic pain genetic variants across open chromatin regions of different cell populations. For the brain snATAC-seq data^36^, we analyzed the following cell types: astrocytes, hippocampal excitatory neurons, isocortical astrocytes, isocortical excitatory neurons, isocortical inhibitory neurons, microglia, neurons, nigral astrocytes, nigral neurons, nigral OPC, striatal astrocytes, and striatal inhibitory neurons. For mouse snATAC-seq data^24^, we analyzed the conserved dorsal horn glial and neuronal subtypes. Because of their complexity, multi-allelic variants and those from the major histocompatibility complex (MHC) region were excluded. Cell types were considered to be significantly associated with chronic pain using an FDR-adjusted *p*-value < 0.05 (treating brain and spinal snATAC-seq or snRNAseq datasets separately).

Following previous examples^82,83^, neurotransmitters for the brain snRNAseq cell clusters were named based on annotations from Siletti et al^30^. Brain regions were assigned to each cell type according to human brain dissections from the same study. Because cell types are not exclusively derived from a single brain region, we assigned a brain region label to a cell cluster only when most of its cells (>50%, typically much higher) originated from that region. We evaluated the enrichment of significant brain cell types in their neurotransmitters and regions using a hypergeometric test.

### MAGMA enrichment analyses

We used MAGMA^37^ in FUMA (v1.3.6a)^95^ to map chronic pain GWAS SNPs to 19,437 protein-coding genes according to their physical position in NCBI build 37. Genes with nonunique names were removed, as were those in the MHC region (chromosome 6, base positions 25,000,000–34,000,000), where the high linkage disequilibrium (LD) among SNPs creates uncertainty as to which mapped genes account for the observed associations. Cell-type analysis in MAGMA v.1.08^37,41^ was applied to test for gene enrichment conditioning on the dataset’s average per-gene expression. We used an FDR-adjusted *p*-value < 0.05 to account for multiple testing and identify significantly enriched cell types (or tissues).

### Gene prioritization for chronic pain across cell types

We used the LDAK-GBAT package^52^ to prioritize genes that contribute significantly to the heritability of chronic pain. To ensure matching of the population LD structure, we used LD reference panels for EUR individuals from 10,000 unrelated UKB participants using LDAK version 5^52^. Exonic and intronic SNPs were mapped to genes based on RefSeq gene annotations^96^. We used the default heritability model in LDAK-GBAT to estimate the heritability attributable to individual genes. We determined the significance of heritable genes for chronic pain using an FDR-adjusted *p*-value < 0.05.

Next, we used fGSEA^53,97^ to identify brain and hDRG cell types with the top-most enriched heritable gene sets. To annotate pathways for chronic pain, we used the gene sets for each enriched cell type as input for ToppGene^54^ to test for over-representation for GO: Molecular Functions, GO: Biological Processes, and GO: Cellular Components. Gene sets and pathways with an FDR-corrected *p*-value < 0.05 were considered significant.

### DATA AVAILABILITY

The full summary statistics from the GWAS meta-analyses will be available upon request. Each of the summary statistics included in the meta-analyses and the multi-omics datasets included in the enrichment analyses are publicly available.

### AUTHOR CONTRIBUTIONS

Project conception: H.R.K and S.T. Study design and data collection: S.T., M.P., M.J.L., C.S., H.Y., A.A-T., U.F-E., C.H., M.C., W.L., A.R.P., R.P.S., R.L.K., T.J.P., and H.R.K. Data analysis: S.T., M.P., M.J.L., and C.S. Study supervision: H.R.K., A.R.P., R.P.S., R.L.K., T.J.P., and L.D. Writing the manuscript: S.T., M.P., M.J.L., C.S., T.J.P., S.G.W., and H.R.K. All authors reviewed and approved the final version of the manuscript.

## ACKNOWLEDGEMENTS

This work was supported by the US Department of Veterans Affairs (grants I50 RX002999-01 to S.G.W., I01 BX003341 to H.R.K. and the VISN 4 Mental Illness Research, Education and Clinical Center) and NIH (grants K99 DA060906 to S.T., F31 NS134318 to M.J.L., U19 NS135528 to W.L., R56 NS133364 to A.R.P. and R.P.S, K01 AA028292 to R.L.K., U19 NS130608 to T.J.P, M.C., and C.P.H, and P30 DA046345 to H.R.K.). The funders had no role in study design, data collection and analysis, decision to publish or preparation of the manuscript. The views expressed in this article are those of the authors and do not necessarily represent the position or policy of the Department of Veterans Affairs or the US Government.

## REFERENCES

1. Nicholas, M. et al. The IASP classification of chronic pain for ICD-11: chronic primary pain. PAIN 160, (2019).

2. Cohen, S. P., Vase, L. & Hooten, W. M. Chronic pain: an update on burden, best practices, and new advances. The Lancet 397, 2082–2097 (2021).

3. Ray, B. M., Kelleran, K. J., Fodero, J. G. & Harvell-Bowman, L. A. Examining the Relationship Between Chronic Pain and Mortality in U.S. Adults. J. Pain 25, (2024).

4. Maher, D. P., Wong, C. H., Siah, K. W. & Lo, A. W. Estimates of Probabilities of Successful Development of Pain Medications: An Analysis of Pharmaceutical Clinical Development Programs from 2000 to 2020. Anesthesiology 137, (2022).

5. Els, C. et al. Adverse events associated with medium- and long-term use of opioids for chronic non-cancer pain: an overview of Cochrane Reviews. Cochrane Database Syst. Rev. (2017) doi:10.1002/14651858.CD012509.pub2.

6. Soliman, N. et al. Pharmacotherapy and non-invasive neuromodulation for neuropathic pain: a systematic review and meta-analysis. Lancet Neurol. 24, 413–428 (2025).

7. Zeng, X., Powell, R. & Woolf, C. J. Mechanism-based nonopioid analgesic targets. J. Clin. Invest. 135, (2025).

8. Minikel, E. V., Painter, J. L., Dong, C. C. & Nelson, M. R. Refining the impact of genetic evidence on clinical success. Nature 629, 624–629 (2024).

9. Maixner, W., Fillingim, R. B., Williams, D. A., Smith, S. B. & Slade, G. D. Overlapping Chronic Pain Conditions: Implications for Diagnosis and Classification. J. Pain 17, T93– T107 (2016).

10. Zorina-Lichtenwalter, K. et al. Genetic risk shared across 24 chronic pain conditions: identification and characterization with genomic structural equation modeling. PAIN 164, (2023).

11. Tanguay-Sabourin, C. et al. A prognostic risk score for development and spread of chronic pain. Nat. Med. 29, 1821–1831 (2023).

12. Apkarian, A. V., Baliki, M. N. & Geha, P. Y. Towards a theory of chronic pain. Prog. Neurobiol. 87, 81–97 (2009).

13. Barroso, J., Branco, P. & Apkarian, A. V. Brain mechanisms of chronic pain: critical role of translational approach. Transl. Res. 238, 76–89 (2021).

14. Johnston, K. J. A. et al. Genome-wide association study of multisite chronic pain in UK Biobank. PLOS Genet. 15, e1008164 (2019).

15. Mocci, E. et al. Genome wide association joint analysis reveals 99 risk loci for pain susceptibility and pleiotropic relationships with psychiatric, metabolic, and immunological traits. PLoS Genet. 19, e1010977 (2023).

16. Toikumo, S. et al. A multi-ancestry genetic study of pain intensity in 598,339 veterans. Nat. Med. 30, 1075–1084 (2024).

17. Khoury, S. et al. Genome-wide analysis identifies impaired axonogenesis in chronic overlapping pain conditions. Brain 145, 1111–1123 (2022).

18. Parisien, M. et al. Sex-specific genetics underlie increased chronic pain risk in women: genome-wide association studies from the UK Biobank. Br. J. Anaesth. doi:10.1016/j.bja.2025.04.013.

19. Bortsov, A. V. et al. Brain-specific genes contribute to chronic but not to acute back pain. PAIN Rep. 7, (2022).

20. Todd, A. J. Neuronal circuitry for pain processing in the dorsal horn. Nat. Rev. Neurosci. 11, 823–836 (2010).

21. Tansley, S. N., Wong, C., Uttam, S., Mogil, J. S. & Khoutorsky, A. Translation regulation in the spinal dorsal horn – A key mechanism for development of chronic pain. Transl. Regul. Pain 4, 20–26 (2018).

22. Hodge, R. D. et al. Conserved cell types with divergent features in human versus mouse cortex. Nature 573, 61–68 (2019).

23. Yadav, A. et al. A cellular taxonomy of the adult human spinal cord. Neuron 111, 328–344.e7 (2023).

24. Arokiaraj, C. M. et al. Spatial, transcriptomic, and epigenomic analyses link dorsal horn neurons to chronic pain genetic predisposition. Cell Rep. 43, (2024).

25. Toikumo, S. et al. Gene discovery and pleiotropic architecture of Chronic Pain in a Genome-wide Association Study of >1.2 million Individuals. medRxiv 2025.02.28.25323112 (2025) doi:10.1101/2025.02.28.25323112.

26. Chen, C. et al. Neural circuit basis of placebo pain relief. Nature 632, 1092–1100 (2024).

27. Kiritoshi, T. et al. Cells and circuits for amygdala neuroplasticity in the transition to chronic pain. Cell Rep. 43, (2024).

28. Mercer Lindsay, N., Chen, C., Gilam, G., Mackey, S. & Scherrer, G. Brain circuits for pain and its treatment. Sci. Transl. Med. 13, eabj7360.

29. Middleton, S. J. et al. Studying human nociceptors: from fundamentals to clinic. Brain 144, 1312–1335 (2021).

30. Siletti, K. et al. Transcriptomic diversity of cell types across the adult human brain. Science 382, eadd7046.

31. Yu, H. et al. Leveraging deep single-soma RNA sequencing to explore the neural basis of human somatosensation. Nat. Neurosci. 27, 2326–2340 (2024).

32. Zeisel, A. et al. Molecular Architecture of the Mouse Nervous System. Cell 174, 999–1014.e22 (2018).

33. Finucane, H. K. et al. Partitioning heritability by functional annotation using genome-wide association summary statistics. Nat. Genet. 47, 1228–1235 (2015).

34. Dib-Hajj, S. D., Cummins, T. R., Black, J. A. & Waxman, S. G. Sodium Channels in Normal and Pathological Pain. Annual Review of Neuroscience vol. 33 325–347 (2010).

35. Harlow, C. E. et al. GWAS of Extended Prescription Analgesic Use Identifies Novel Genetic Loci in Chronic Pain. medRxiv 2024.12.02.24318312 (2024) doi:10.1101/2024.12.02.24318312.

36. Corces, M. R. et al. Single-cell epigenomic analyses implicate candidate causal variants at inherited risk loci for Alzheimer’s and Parkinson’s diseases. Nat. Genet. 52, 1158–1168 (2020).

37. de Leeuw, C. A., Mooij, J. M., Heskes, T. & Posthuma, D. MAGMA: Generalized Gene-Set Analysis of GWAS Data. PLOS Comput. Biol. 11, e1004219 (2015).

38. Woo, C.-W. et al. Quantifying cerebral contributions to pain beyond nociception. Nat. Commun. 8, 14211 (2017).

39. Tu, Y. et al. Pain-preferential thalamocortical neural dynamics across species. *Nat*. Hum. Behav. 8, 149–163 (2024).

40. Skene, N. G. et al. Genetic identification of brain cell types underlying schizophrenia. Nat. Genet. 50, 825–833 (2018).

41. Watanabe, K., Umićević Mirkov, M., de Leeuw, C. A., van den Heuvel, M. P. & Posthuma, D. Genetic mapping of cell type specificity for complex traits. Nat. Commun. 10, 3222 (2019).

42. Pan, T. et al. Glutamatergic neurons and myeloid cells in the anterior cingulate cortex mediate secondary hyperalgesia in chronic joint inflammatory pain. Brain. Behav. Immun. 101, 62–77 (2022).

43. Coppieters, I., Cagnie, B., De Pauw, R., Meeus, M. & Timmers, I. Enhanced amygdala-frontal operculum functional connectivity during rest in women with chronic neck pain: Associations with impaired conditioned pain modulation. NeuroImage Clin. 30, 102638 (2021).

44. Gudin, J., Sakr, M., Fason, J. & Hurwitz, P. Piezo Ion Channels and Their Association With Haptic Technology Use: A Narrative Review. Cureus 17, e77433 (2025).

45. Gautam, M. et al. Distinct local and global functions of mouse Aβ low-threshold mechanoreceptors in mechanical nociception. Nat. Commun. 15, 2911 (2024).

46. Yang, L. et al. Human and mouse trigeminal ganglia cell atlas implicates multiple cell types in migraine. Neuron 110, 1806–1821.e8 (2022).

47. Bhuiyan, S. A. et al. Harmonized cross-species cell atlases of trigeminal and dorsal root ganglia. Sci. Adv. 10, eadj9173.

48. Mogil, J. S., Parisien, M., Esfahani, S. J. & Diatchenko, L. Sex differences in mechanisms of pain hypersensitivity. Neurosci. Biobehav. Rev. 163, 105749 (2024).

49. Ray, P. R. et al. RNA profiling of human dorsal root ganglia reveals sex differences in mechanisms promoting neuropathic pain. Brain 146, 749–766 (2023).

50. Tavares-Ferreira, D. et al. Spatial transcriptomics of dorsal root ganglia identifies molecular signatures of human nociceptors. Sci. Transl. Med. 14, eabj8186.

51. Johnston, K. J. A. et al. Sex-stratified genome-wide association study of multisite chronic pain in UK Biobank. PLoS Genet. 17, e1009428 (2021).

52. Berrandou, T.-E., Balding, D. & Speed, D. LDAK-GBAT: Fast and powerful gene-based association testing using summary statistics. Am. J. Hum. Genet. 110, 23–29 (2023).

53. Korotkevich, G. et al. Fast gene set enrichment analysis. bioRxiv 060012 (2021) doi:10.1101/060012.

54. Chen, J., Bardes, E. E., Aronow, B. J. & Jegga, A. G. ToppGene Suite for gene list enrichment analysis and candidate gene prioritization. Nucleic Acids Res. 37, W305–W311 (2009).

55. Berta, T. et al. Gene Expression Profiling of Cutaneous Injured and Non-Injured Nociceptors in SNI Animal Model of Neuropathic Pain. Sci. Rep. 7, 9367 (2017).

56. Jahangiri Esfahani, S., Parisien, M., Surbey, C. & Diatchenko, L. The Transcriptomics Pain Signature Database. *bioRxiv* 2023.06.16.545337 (2023) doi:10.1101/2023.06.16.545337.

57. Parisien, M. et al. Genetic pathway analysis reveals a major role for extracellular matrix organization in inflammatory and neuropathic pain. PAIN 160, (2019).

58. Arendt-Tranholm, A., et al. Single-cell characterization of the human C2 dorsal root ganglion recovered from C1-2 arthrodesis surgery: implications for neck pain. *bioRxiv* 2025.03.24.645122 (2025) doi:10.1101/2025.03.24.645122.

59. Franco-Enzástiga, Ú. et al. Epigenomic landscape of the human dorsal root ganglion: sex differences and transcriptional regulation of nociceptive genes. PAIN 166, (2025).

60. Temmermand, R., Barrett, J. E. & Fontana, A. C. K. Glutamatergic systems in neuropathic pain and emerging non-opioid therapies. Pharmacol. Res. 185, 106492 (2022).

61. Peek, A. L. et al. Brain GABA and glutamate levels across pain conditions: A systematic literature review and meta-analysis of 1H-MRS studies using the MRS-Q quality assessment tool. NeuroImage 210, 116532 (2020).

62. Li, M. & Yang, G. A mesocortical glutamatergic pathway modulates neuropathic pain independent of dopamine co-release. Nat. Commun. 15, 643 (2024).

63. Naylor, B. et al. Reduced Glutamate in the Medial Prefrontal Cortex Is Associated With Emotional and Cognitive Dysregulation in People With Chronic Pain. Front. Neurol. 10, (2019).

64. Wang, X.-Y. et al. A glutamatergic DRN–VTA pathway modulates neuropathic pain and comorbid anhedonia-like behavior in mice. Nat. Commun. 14, 5124 (2023).

65. Alsaloum, M., Vasylyev, D. & Waxman, S. G. Targeting sodium channels: challenges for acute pain and in the leap to chronic pain. Nat. Rev. Neurol. (in press).

66. Dubin, A. & Patapoutian, A. Nociceptors: the sensors of the pain pathway. J Clin Invest 120, 3760–72 (2010).

67. Kupari, J. et al. Single cell transcriptomics of primate sensory neurons identifies cell types associated with chronic pain. Nat. Commun. 12, 1510 (2021).

68. Cavanaugh, D. J. et al. Distinct subsets of unmyelinated primary sensory fibers mediate behavioral responses to noxious thermal and mechanical stimuli. Proc. Natl. Acad. Sci. 106, 9075–9080 (2009).

69. Wercberger, R., Braz, J. M., Weinrich, J. A. & Basbaum, A. I. Pain and itch processing by subpopulations of molecularly diverse spinal and trigeminal projection neurons. Proc. Natl. Acad. Sci. 118, e2105732118 (2021).

70. Kimmey, B. A. et al. Convergent state-control of endogenous opioid analgesia. *bioRxiv* 2025.01.03.631111 (2025) doi:10.1101/2025.01.03.631111.

71. Faber, C. G. et al. Gain of function NaV1.7 mutations in idiopathic small fiber neuropathy. Ann. Neurol. 71, 26–39 (2012).

72. Faber, C. G. et al. Gain-of-function Nav1.8 mutations in painful neuropathy. Proc. Natl. Acad. Sci. 109, 19444–19449 (2012).

73. Huang, J. et al. Gain-of-function mutations in sodium channel NaV1.9 in painful neuropathy. Brain 137, 1627–1642 (2014).

74. Rush, A. M., Cummins, T. R. & Waxman, S. G. Multiple sodium channels and their roles in electrogenesis within dorsal root ganglion neurons. J. Physiol. 579, 1–14 (2007).

75. Dib-Hajj, S., Black, J. A., Cummins, T. R. & Waxman, S. G. NaN/Nav1.9: a sodium channel with unique properties. Trends Neurosci. 25, 253–259 (2002).

76. Herzog, R. I., Cummins, T. R. & Waxman, S. G. Persistent TTX-Resistant Na+ Current Affects Resting Potential and Response to Depolarization in Simulated Spinal Sensory Neurons. J. Neurophysiol. 86, 1351–1364 (2001).

77. Alsaloum, M. et al. Contributions of NaV1.8 and NaV1.9 to excitability in human induced pluripotent stem-cell derived somatosensory neurons. Sci. Rep. 11, 24283 (2021).

78. Rush, A. M. & Waxman, S. G. PGE2 increases the tetrodotoxin-resistant Nav1.9 sodium current in mouse DRG neurons via G-proteins. Brain Res. 1023, 264–271 (2004).

79. Wang, L. et al. Molecular and cellular dynamics of the developing human neocortex. Nature (2025) doi:10.1038/s41586-024-08351-7.

80. Lu, T. et al. Decoding transcriptional identity in developing human sensory neurons and organoid modeling. Cell 187, 7374–7393.e28 (2024).

81. Finucane, H. K. et al. Heritability enrichment of specifically expressed genes identifies disease-relevant tissues and cell types. Nat. Genet. 50, 621–629 (2018).

82. Yao, S. et al. Connecting genomic results for psychiatric disorders to human brain cell types and regions reveals convergence with functional connectivity. Nat. Commun. 16, 395 (2025).

83. Duncan, L. E. et al. Mapping the cellular etiology of schizophrenia and complex brain phenotypes. Nat. Neurosci. 28, 248–258 (2025).

84. Farrar, J. T. A consideration of differences in pain scales used in clinical trials. PAIN 163, (2022).

85. Euasobhon, P. et al. Reliability and responsivity of pain intensity scales in individuals with chronic pain. PAIN 163, (2022).

86. Kurki, M. I. et al. FinnGen provides genetic insights from a well-phenotyped isolated population. Nature 613, 508–518 (2023).

87. Yengo, L. et al. A saturated map of common genetic variants associated with human height. Nature 610, 704–712 (2022).

88. Locke, A. E. et al. Genetic studies of body mass index yield new insights for obesity biology. Nature 518, 197–206 (2015).

89. Thul, P. J. & Lindskog, C. The human protein atlas: A spatial map of the human proteome. Protein Sci. 27, 233–244 (2018).

90. The GTEx Consortium et al. The GTEx Consortium atlas of genetic regulatory effects across human tissues. Science 369, 1318–1330 (2020).

91. Kawaji, H., Kasukawa, T., Forrest, A., Carninci, P. & Hayashizaki, Y. The FANTOM5 collection, a data series underpinning mammalian transcriptome atlases in diverse cell types. Sci. Data 4, 170113 (2017).

92. Zhong, W., et al. The neuropeptide landscape of human prefrontal cortex. Proc. Natl. Acad. Sci. 119, e2123146119 (2022).

93. Monaco, G. et al. RNA-Seq Signatures Normalized by mRNA Abundance Allow Absolute Deconvolution of Human Immune Cell Types. Cell Rep. 26, 1627–1640.e7 (2019).

94. Srinivasan, C. et al. Addiction-Associated Genetic Variants Implicate Brain Cell Type- and Region-Specific Cis-Regulatory Elements in Addiction Neurobiology. J. Neurosci. 41, 9008 (2021).

95. Watanabe, K., Taskesen, E., van Bochoven, A. & Posthuma, D. Functional mapping and annotation of genetic associations with FUMA. Nat. Commun. 8, 1826 (2017).

96. O’Leary, N. A. et al. Reference sequence (RefSeq) database at NCBI: current status, taxonomic expansion, and functional annotation. Nucleic Acids Res. 44, D733–D745 (2016).

97. Subramanian, A. et al. Gene set enrichment analysis: A knowledge-based approach for interpreting genome-wide expression profiles. Proc Natl Acad Sci USA 102, 15545–15550 (2005).

