## Supplemental Figures 1-3 for "Etiological basis for chronic pain genetic variation in brain and dorsal root ganglia cell types"

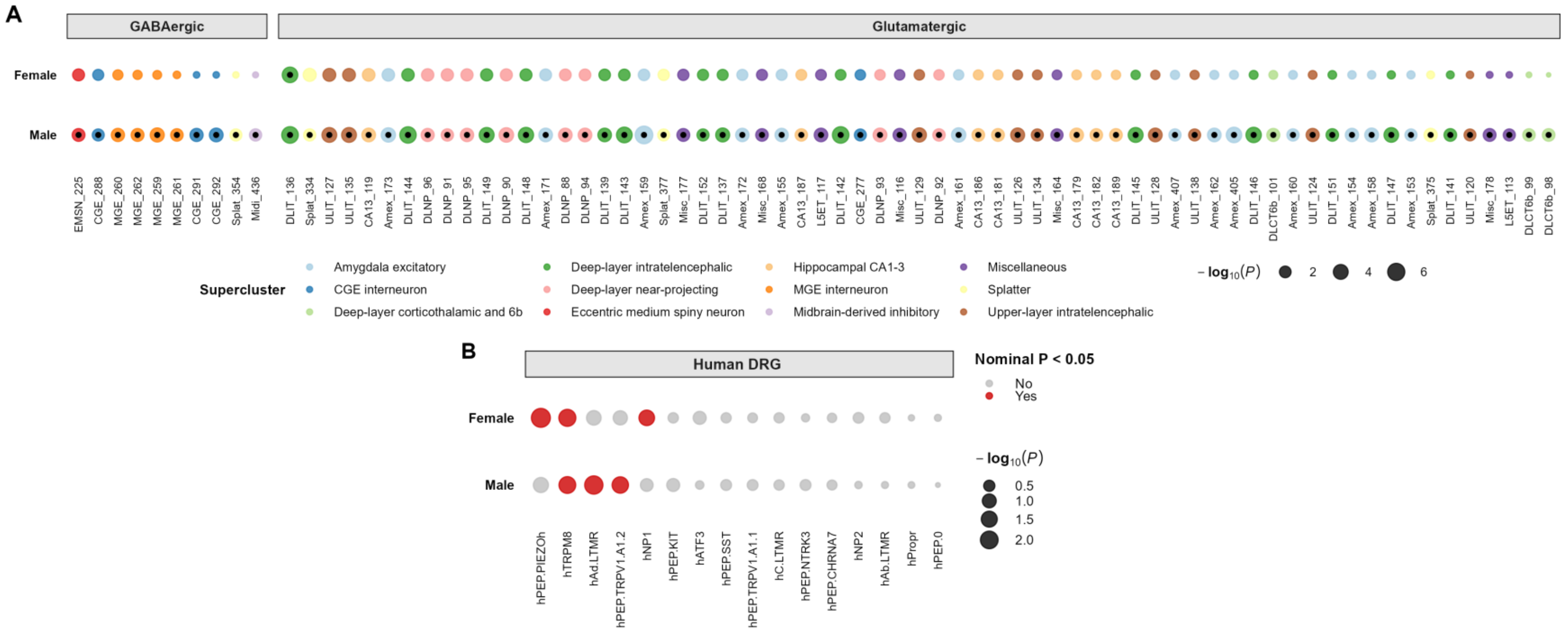


**Supplemental Figure 1. Sex-stratified S-LDSC cell type enrichment analysis**. **(A)** The significantly enriched cell types across sex (females, $n = 1$; males, $n = 75$; FDR $p < 0.05$) among 461 brain cell clusters. Each cell type is colored based on its supercluster, and cell types are grouped into GABAergic ($n = 10$) and glutamatergic ($n = 75$) based on their neurotransmitter annotation. Cell types that are significantly associated are marked by a black dot centered within each colored circle. In total, 1 significant brain cell cluster (DLIT_136) overlaps across sexes. **(B)** Human single-soma DRG cell type enrichment. Cell type enrichments denoted by red in **B** ($n = 5$) are nominally significantly enriched for chronic pain ($P < 0.05$). One hDRG cell type (hTRPM8) overlaps across sexes. See Supplemental Tables 11 and 12 for full results.

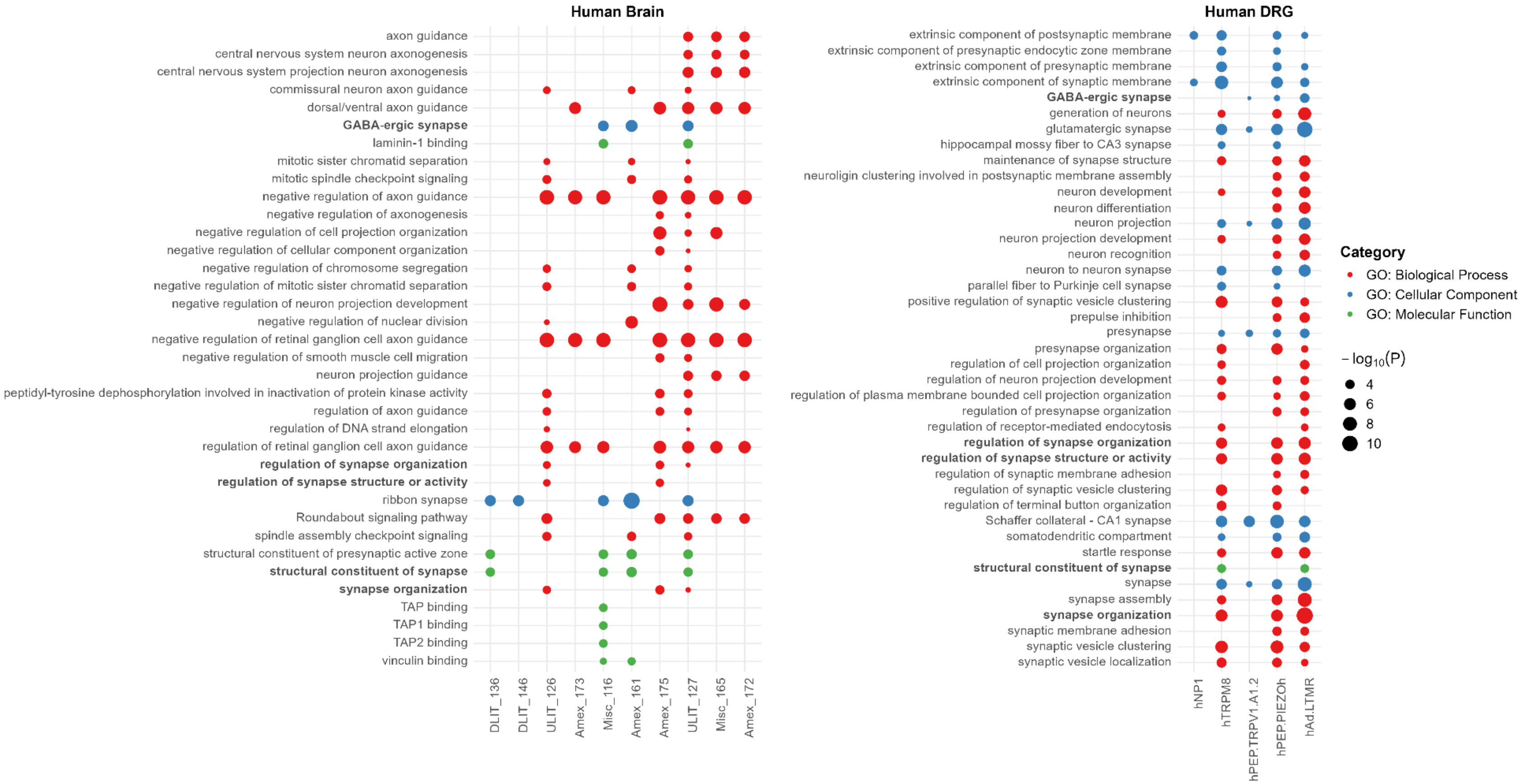


**Supplemental Figure 2. Enriched GO categories in brain and hDRG cell types.** Top enriched GO categories among 10 and 5 cell clusters in brain and hDRG, respectively, are shown. Each dot is colored based on GO categories. Overlapping GO categories in brain and hDRG are annotated in bold. See Supplemental Tables 16 and 17 for full results.

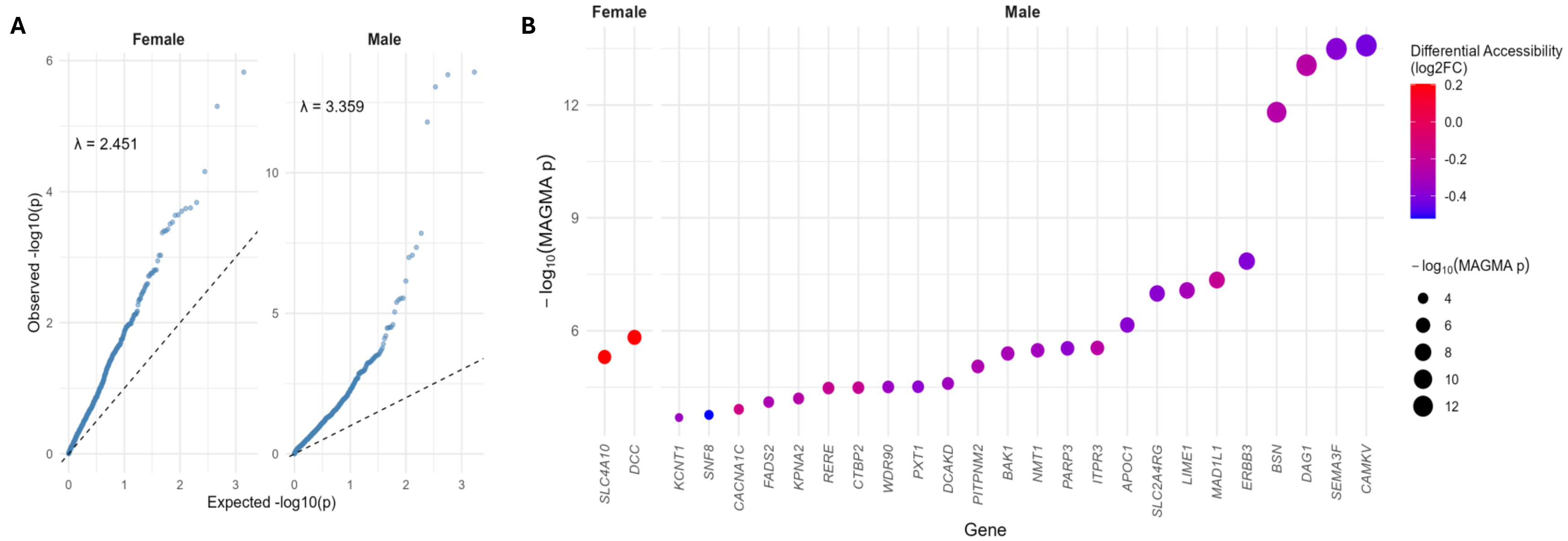


**Supplemental Figure 3. Sex-specific chronic pain associated genes in human DRG differentially accessible regions**. **(A)** QQ plots of pain-associated genes in sex-differential chromatin accessible hDRG neurons. **(B)** Top differentially accessible genes in neuronal hDRG neurons associated with chronic pain risk in females and males. See Supplemental Table 19 for details
